# Cooking fuel and symptoms of chronic respiratory disease in ageing adults: Evidence from West Africa and North America

**DOI:** 10.1101/2024.04.25.24306374

**Authors:** Gabriel Dusing, Eyram Adzo Agbe, Reginald Quansah, Godfred O. Boateng

## Abstract

**Background:** The combustion of solid fuels (e.g., wood, coal, and charcoal) for cooking fuel is widespread in low– and middle-income countries. The use of these fuels increases exposure to airborne pollutants which have been shown to increase the risk of disease and premature death, particularly among older individuals. Despite this, most studies examining this association come from India and China. Furthermore, few studies have examined this association among the elderly. This study aims to fill this gap by examining the association between household cooking fuel and chronic respiratory disease.

**Methods:** We analyzed data from Wave 2 of the World Health Organization’s Study on Global Ageing and Adult Health. Our study sample consists of 7,253 adults aged 50+ from Mexico and Ghana. Logistic regression was conducted to study the association between chronic respiratory disease (measured by whether the participant reported having either (1) a medical diagnosis for chronic obstructive pulmonary disorder, chronic bronchitis, or emphysema, or (2) symptoms suggesting chronic lung infection), and the main cooking fuel used by the household.

**Results:** The prevalence of chronic respiratory disease was 6.55% and 17.45% for Ghana and Mexico, respectively. 15.96%, and 22.47% of participants lived in households using solid fuels for cooking. Adjusting for covariates, solid fuel use was associated with 1.72 (95%CI: 1.07-2.79; p=0.026) times higher odds of chronic respiratory disease. Regional disparities were found, with individuals in Mexico and Ghana respectively having 1.70 (95%CI:1.01-2.89; p=0.049) and 3.40 (95%CI:1.50-7.72; p=0.004) higher adjusted odds for chronic respiratory disease.

**Conclusions:** Our results confirm the association found between the use of solid fuels and increased chronic respiratory disease risk. These findings demonstrate the salience of improving access to clean cooking fuels and methods, especially for households in the sub-Saharan region, particularly, women. Policy interventions need to capture the unique needs of women to ensure that health outcomes from energy use are well-mitigated.

## Introduction

In 2017 alone, chronic respiratory diseases (CRDs) were responsible for almost 4 million deaths globally and a further 1,470 disability life-adjusted years (DALYs) per 100,000. The World Health Organizations (WHO) classifies CRDs as a group of diseases that affect the airways and the lungs, including diseases such as asthma, chronic obstructive pulmonary disorder (COPD), and pulmonary hypertension (1,2). They are characterized by symptoms that include persistent cough, wheezing, dyspnea, and sputum production (3,4). CRDs are also further responsible for major economic losses; in the United States, for example, the mean annual indirect costs of COPD alone were estimated to range between US$1,525-$3,340 per person in 2010 dollars (5). However, the economic and societal burden of CRDs are borne disproportionately by low– and middle-income countries (LMICs) (6,7).

The WHO has listed COPD and asthma as the most common CRDs (8). COPD is a chronic inflammatory disease of the lungs having an estimated global prevalence between 11.7% (9) and 12.2% (10). Both studies producing these estimates found that the Americas had the higher prevalence (approximately 15%) compared to Africa (about 10%), but both studies attribute this disparity to the absence of studies examining COPD in the latter region. A 2015 review further attributed this disparity to lower awareness and subsequent underdiagnosis of COPD in these regions (11), which was shown to be a persistent issue as recently as 2019 (12–14). Asthma is a CRD characterized by episodes of wheezing, coughing, shortness of breath, and chest tightness, triggered by factors such as strenuous physical activity or exposure to airborne pollutants (15,16). While asthma is typically diagnosed in early age, a diagnosis can occur at any stage in life (15), with adult-onset asthma associated with moderate to severe symptoms (17,18). A review of over 200 population-based studies from over 80 countries estimated the global prevalence of asthma in 2019 was 9.8% (19). This review also found that the African region had the highest prevalence among individuals aged 5-69 at 11.3%, and had the highest prevalence of current wheezing symptoms 13.2% (compared to 11.5% globally). While COPD and asthma are CRDs that have received the most attention, other diseases such as occupational lung disease (20) and pulmonary hypertension (21) are no less debilitating, but have been shown by prior research to be underdiagnosed, especially among older adults.

The considerable prevalence of CRDs worldwide masks the equally consequential impact of their symptoms on the quality of life especially on older adults. A 2019 longitudinal study of 1,000 Korean adults (age 65+) found that chronic cough was associated with over 3 times higher odds of having a depression diagnosis, measured through the Geriatric Depression Scale (GDS) and the Center for Epidemiologic Studies Depression Scale (CES-D) (22). A 2016 study of 887 Portuguese older adults found that reporting wheezing in the past 12-months was associated with 2.03 higher odds reporting lower quality of life, as measured by the World Health Organization QOL questionnaire (23). These findings suggest that the impact of CRD symptoms on quality of life can be as critical, if not more so, than the clinical diagnosis of the disease itself.

Lifestyle factors have been shown to be significant risk factors for CRDs. For example, current cigarette smokers have consistently been shown to have higher risk for both diagnosis and symptoms of CRDs (24–26). Environmental factors, such as pollution, have also been shown to contribute to significantly higher CRD risk. For example, among 2,397 elderly adults from South Africa, individuals living within a 2 km radius of a mine dump had 1.57, 2.02, and 2.01 higher odds respectively of asthma, chronic cough, and wheezing compared to those living 5 km away or more (27). A large-scale cohort study in Ontario with 5.1 million adults aged 35-85 and utilizing linked administrative data in Ontario found that each unit increase of common pollutants – PM2.5, NOx, and O3 – was respectively associated with 1.07, 1.04, and 1.04 times higher risk of developing COPD. This association was based on exposure levels calculated as a three-year moving average (28). Exposure to these airborne pollutants and heightened CRD risk is thought to be driven by inflammation. A 2014 study found that exposure to ambient air pollution resulted in increases in biomarkers of inflammation, specifically, a 51% increase c-reactive protein, and 12% increase in interleukin-6. Prolonged exposure to these pollutants, and subsequent inflammation, have been shown to be associated with potentially irreversible changes to lung tissue (29,30).

The correlation between environmental pollution and CRD risk motivates the examination of household air pollution on CRDs given the overlap of pollutants in both of these contexts (31–33). Furthermore, the widespread practice of the combustion of solid fuels (such as, wood, coal, charcoal) and kerosene in many LMICs is a major source of household air pollution (31,34,35) and has been linked to heightened risk of CRDs in these regions (35–39). The majority of the extant literature examining this association is based on studies conducted in China and India, with fewer studies from other LMICs in regions such as sub-Saharan Africa or Central America. This gap underscores the importance of examining this association in countries like Ghana with its high prevalence of solid fuel use. A 2015 study of 8,267 Ghanaian households found that less than 10% of participants reported using liquified natural gas (LNG) or electricity to cook (40). This study additionally found that over 57% of respondents cooked with firewood and almost 30% with charcoal. In contrast, a study of 14,245 adults aged 50+ in Mexico found that while the overwhelming majority (81.75%) of respondents cooked exclusively with LNG, about 10% of respondents cooked primarily or exclusively used solid fuels such as wood (41). These contrasting fuel usage patterns in Ghana and Mexico provide a unique comparative perspective to explore the impact of different cooking fuels on the prevalence and severity of CRD symptoms. Policymakers can use this information to develop targeted interventions, such as promoting cleaner cooking fuels and improving ventilation in homes, in these contexts.

This study utilizes a population-representative sample of adults aged 50 and above from two countries in West Africa (Ghana) and North America (Mexico) respectively to investigate the relationship between the type of cooking fuel used and the prevalence of symptoms associated with chronic lung diseases (CLDs). Drawing on existing research, we hypothesize that individuals residing in households where solid fuels and kerosene are the primary cooking fuels have a higher likelihood of exhibiting symptoms indicative of CLDs. This hypothesis is grounded in the understanding that exposure to emissions from these fuels may contribute significantly to the development or exacerbation of respiratory conditions.

## Materials and methods

### Study design and participants

We conducted a cross-sectional analysis of Wave 2 (2014–2015) from the Study on Global AGEing and Adult Health (SAGE), pooling samples from Ghana and Mexico. The SAGE is an ongoing, WHO-initiated study examining health and other characteristics among older adults from 6 participating countries including Ghana and Mexico (42). Wave 2 is the most currently available (as of the time of writing) data from Mexico and Ghana. The survey was conducted by trained interviewers, and the representativeness of the sample to each country’s social, economic, and demographic constitution was ensured through multistage cluster sampling and subsequent sampling weights computed for each individual. To ensure generalizability, all analyses in this study were conducted with the application of these weights.

### Ethics and permissions

Data from SAGE Ghana wave 2 and SAGE Mexico wave 2 were used for this study. WHO Ethics Review Committee (reference number RPC149) with local approval from the University of Ghana Medical School Ethics and Protocol Review Committee. The Ethics and Research Committees of the National Institute of Public Health in Mexico approved the Mexico SAGE data. All participants were provided with a detailed explanation of study procedures and signed an informed consent letter. The necessary permission was obtained from the World Health Organization to use these data.

### Outcome

The outcome of this study was having symptoms or medical diagnosis of a CRD. This was determined through four questions in the study, through which respondents were given the option of answering yes or no: [1] “Have you ever been diagnosed with chronic respiratory disease (emphysema, bronchitis, COPD)?”, [2] “During the last 12 months, have you experienced any shortness of breath at rest? (while awake)”, [3] “During the last 12 months, have you experienced any coughing or wheezing for ten minutes or more at a time?”, [4] “During the last 12 months, have you experienced any coughing up sputum or phlegm for most days of the month for at least 3 months?”. Respondents were considered to have a chronic respiratory disease if they responded “yes” to at least one of questions [1] – [4], and in contrast, were considered to not have a chronic respiratory disease if they responded “no” to all of questions [1] – [4]. This is similar to other studies that have examined chronic respiratory disease using SAGE (43). The use of symptoms as indicators of CRDs, rather than spirometry readings, was informed by evidence from prior studies indicating that individuals may still experience debilitating respiratory symptoms, even with normal spirometry measurements (44,45). Additionally, the presence of these symptoms has been demonstrated to be highly predictive of future mortality and morbidity (46), lending further credibility to their importance in assessing CRDs.

### Exposure

The exposure of interest in this study was the type of cooking fuel primarily used in the respondent’s home through the question “What type of fuel does your household mainly use for cooking?” Respondents could choose from options including gas, electricity, kerosene/paraffin, or solid fuels and biomass such as coal/charcoal, animal dung, wood, or other plant matter. The responses were then categorized into two groups for analysis: one group for those using gas or electricity, and another group comprising all other options.

### Covariates

Multivariate models included adjustments for the following variables: sex, age, rural-urban status, education, waist-to-hip ratio, and smoking status. In addition to socio-demographic characteristics (sex and age), rural/urban status and education were included as measures of socio-economic characteristics. Smoking status (current, former, non-smoker, and not stated) was included as both a physical health indicator (47–50) and its role as a major risk factor for CRDs (51,52). Continuous waist-to-hip ratio was included as an additional physical health indicator (53,54).

Adjusted models also included adjustments for cooking space to account for differences in exposure. Respondents who indicated that they cooked with solid fuels were further asked: “Where is cooking usually done?”, with possible responses 1 “In a room used for living or sleeping”, 2 “In a separate room used as kitchen”, 3 “In a separate building used as kitchen”, 4 “Outdoor”, or 7 “Other, specify:”. This covariate was coded into a binary category where respondents who indicated that they cooked in the same room used for living or sleeping were included in one category, and other responses in the other category.

### Statistical analyses

A series of logistic regression analyses to explore the relationship between primary household cooking fuel and the odds of CRD. These analyses were conducted using both unadjusted and adjusted models, with each analysis stratified by country. Adjusted models were further stratified by sex, recognizing the historically greater exposure to cooking fuels that women experience due to traditional gender roles in Ghana and Mexico.

As part of our preliminary analysis, we calculated the Intraclass Correlation Coefficient (ICC) to assess the degree of clustering of CRDs within countries. The ICC quantifies the proportion of total variance in the outcome (reporting symptoms of CRDs) that is attributable to differences between groups at the country level. The ICC was calculated using a null model — a multilevel logistic regression model with no predictors except for the random intercepts for countries. This model partitions the variance into within-country and between-country components. We considered an ICC value greater than 0.40 as indicative of sufficient between-country variance to warrant multilevel modeling based on prior literature (55,56).

In our analysis, descriptive tables were generated to illustrate the distribution of sample characteristics, stratified both by the outcome status and by country. We evaluated the bivariate associations between each characteristic and the outcome using the chi-square test for categorical variables and the F-test for continuous variables. The threshold for statistical significance was set at a 5% confidence level across all analyses. All statistical procedures were conducted using STATA 18 (57).

## Results

### Sample characteristics

Table 1 describes our study sample by CRD status, with further detailed breakdown by country. Our overall study sample consists of 7,243 adults aged 50+ from Mexico and Ghana weighted to represent 23,073,224 individuals. For brevity, the sample characteristics for the sample combining Mexico and Ghana are presented in in S1 table, and only the breakdown by country are presented in our main results. Table 1 presents unweighted counts for transparency, but proportions and p-values were calculated using weighted values to ensure representativeness to their respective countries. The overall prevalence of CRDs was 15.96%. By country, the prevalence of CRDs in Ghana was 6.55%, but this rate was almost triple in Mexico at 17.54%.

**Table 1.** Sample characteristics chronic respiratory disease status, with further breakdown by country (unweighted N=7,243; weighted N=23,073,224)

| Mexico and Ghana |  |  |  |  |  |  |  |
| --- | --- | --- | --- | --- | --- | --- | --- |
|  | No Diagnosis or Symptoms of<br>chronic respiratory disease |  | Has Diagnosis or Symptoms of<br>chronic respiratory disease |  | Total |  | p-values |
|  | Unweighted<br>Counts | Weighted Row % | Unweighted<br>Counts | Weighted Row % | Unweighted<br>Counts | Weighted Row % |  |
| **Totals** | 6,346 | 84.04% | 897 | 15.96% | 7,243 | 100.00% |  |
|  | Unweighted<br>Counts | Weighted Col % | Unweighted<br>Counts | Weighted Col % | Unweighted<br>Counts | Weighted Col % |  |
| **Cooking Fuel** |  |  |  |  |  |  | 0.0008 |
| Wood/Charcoal/C<br>oal/Kerosene | 3,184 | 23.71% | 311 | 15.75% | 3,495 | 22.44% |  |
| Gas/Electricity | 3,162 | 76.29% | 586 | 84.25% | 3,748 | 77.56% |  |
| Ghana Only |  |  |  |  |  |  |  |
|  | No Diagnosis or Symptoms of<br>chronic respiratory disease |  | Has Diagnosis or Symptoms of<br>chronic respiratory disease |  | Total |  | p-values |

|  | Unweighted<br>Counts | Weighted Row % | Unweighted<br>Counts | Weighted Row % | Unweighted<br>Counts | Weighted Row % |  |
| --- | --- | --- | --- | --- | --- | --- | --- |
| **Totals** | 3,092 | 93.45% | 248 | 6.55% | 3,340 | 100.00% |  |
|  | Unweighted<br>Counts | Weighted Col % | Unweighted<br>Counts | Weighted Col % | Unweighted<br>Counts | Weighted Row % |  |
| **Cooking Fuel** |  |  |  |  |  |  | 0.0005 |
| Wood/Charcoal/C<br>oal/Kerosene | 2,773 | 86.85% | 235 | 95.08% | 3,008 | 87.39% |  |
| Gas/Electricity | 319 | 13.15% | 13 | 4.92% | 332 | 12.61% |  |
| **Cooking Space** |  |  |  |  |  |  | 0.0033 |
| Shared with<br>Living/Sleeping<br>Space | 18 | 0.58% | 7 | 2.55% | 25 | 0.71% |  |
| Separate from<br>Living/Sleeping<br>Space | 3,074 | 99.42% | 241 | 97.45% | 3,315 | 99.29% |  |
| **Sex** |  |  |  |  |  |  | 0.8808 |
| Male | 1,291 | 47.21% | 105 | 46.54% | 1,396 | 47.16% |  |
| Female | 1,801 | 52.79% | 143 | 53.46% | 1,944 | 52.84% |  |
| **Age** |  |  |  |  |  |  | p<0.0001 |
| Younger than 65 | 1,731 | 66.72% | 102 | 50.09% | 1,833 | 65.63% |  |
| 65 and Older | 1,361 | 33.28% | 146 | 49.91% | 1,507 | 34.37% |  |
| **Education** |  |  |  |  |  |  | 0.0152 |
| No Secondary Education | 769 | 29.23% | 46 | 18.77% | 815 | 28.55% |  |
| Secondary Education or Higher | 812 | 29.83% | 57 | 29.66% | 869 | 29.82% |  |
| Not Stated | 1,511 | 40.94% | 145 | 51.57% | 1,656 | 41.64% |  |
| **Rural-Urban Status** |  |  |  |  |  |  | 0.074 |
| Urban | 1,199 | 48.78% | 84 | 40.67% | 1,283 | 48.25% |  |
| Rural | 1,893 | 51.22% | 164 | 59.33% | 2,057 | 51.75% |  |
| **Smoking Status** |  |  |  |  |  |  | 0.2739 |
| Never Smoked | 2,878 | 91.56% | 223 | 91.14% | 3,101 | 91.53% |  |
| Current Smoker | 121 | 4.31% | 16 | 6.34% | 137 | 4.44% |  |
| Former Smoker | 21 | 0.65% | 4 | 0.75% | 25 | 0.66% |  |
| Not Stated | 72 | 3.48% | 5 | 1.77% | 77 | 3.37% |  |
| **Waist-to-hip Ratio** | *Mean* | *Std Err* | *Mean* | *Std Err* | *Mean* | *Std Err* | 0.345* |
|  | 0.924 | 0.003 | 0.927 | 0.005 | 0.924 | 0.002 |  |
| **Mexico Only** |  |  |  |  |  |  |  |
|  | **No Diagnosis or Symptoms of chronic respiratory disease** |  | **Has Diagnosis or Symptoms of chronic respiratory disease** |  | **Total** |  | **p-values** |
|  | **Unweighted Counts** | **Weighted Row %** | **Unweighted Counts** | **Weighted Row %** | **Unweighted Counts** | **Weighted Row %** |  |
| **Totals** | 3,254 | 82.46% | 649 | 17.54% | 3,903 | 100.00% |  |
|  | **Unweighted Counts** | **Weighted Col %** | **Unweighted Counts** | **Weighted Col %** | **Unweighted Counts** | **Weighted Row %** |  |
| **Cooking Fuel** |  |  |  |  |  |  | 0.6796 |
| Wood/Charcoal/Coal/Kerosene | 411 | 11.72% | 76 | 10.78% | 487 | 11.55% |  |
| Gas/Electricity | 2,843 | 88.28% | 573 | 89.22% | 3,416 | 88.45% |  |
| **Cooking Space** |  |  |  |  |  |  | 0.0006 |
| Shared with Living/Sleeping Space | 10 | 0.11% | 5 | 0.66% | 15 | 0.21% |  |
| Separate from Living/Sleeping Space | 3244 | 99.89% | 644 | 99.34% | 3888 | 99.79% |  |
| **Sex** |  |  |  |  |  |  | 0.1511 |
| Male | 1,362 | 45.14% | 248 | 40.61% | 1,610 | 44.35% |  |
| Female | 1,892 | 54.86% | 401 | 59.39% | 2,293 | 55.65% |  |
| **Age** |  |  |  |  |  |  | 0.3161 |
| Younger than 65 | 1,419 | 62.20% | 262 | 59.18% | 1,681 | 61.67% |  |
| 65 and Older | 1,835 | 37.80% | 387 | 40.82% | 2,222 | 38.33% |  |
| **Education** |  |  |  |  |  |  | 0.0123 |
| No Secondary Education | 1,905 | 53.95% | 408 | 63.39% | 2,313 | 55.61% |  |
| Secondary Education or Higher | 826 | 32.94% | 141 | 22.27% | 967 | 31.07% |  |
| Not Stated | 523 | 13.10% | 100 | 14.34% | 623 | 13.32% |  |
| **Rural-Urban Status** |  |  |  |  |  |  | 0.1515 |
| Urban | 2,360 | 78.08% | 488 | 82.44% | 2,848 | 78.85% |  |
| Rural | 894 | 21.92% | 161 | 17.56% | 1,055 | 21.15% |  |
| **Smoking Status** |  |  |  |  |  |  | 0.4134 |
| Never Smoked | 2,465 | 73.69% | 476 | 78.13% | 2,941 | 74.47% |  |
| Current Smoker | 300 | 12.37% | 80 | 10.65% | 380 | 12.07% |  |
| Former Smoker | 189 | 5.74% | 31 | 4.19% | 220 | 5.47% |  |
| Not Stated | 300 | 8.20% | 62 | 7.04% | 362 | 8.00% |  |
| **Waist-to-hip**<br>**Ratio** | *Mean* | *Std Err* | *Mean* | *Std Err* | *Mean* | *Std Err* | 0.497* |
|  | 0.983 | 0.013 | 0.970 | 0.006 | 0.981 | 0.011 |  |

\*p-value obtained from F-test.

In Ghana, the overwhelming majority (87.39%) used solid fuels or kerosene as cooking fuels, compared to 12.61% who used gas or electricity. The bivariate association between cooking fuel and CRD status was significant (p=0.0005), and of those who did not have a CRDs, 86.85% used solid fuels or kerosene, while 13.15% used LNG or electricity. In contrast, more (95.08%) people with symptoms or diagnoses used solid fuels or kerosene. The pattern of cooking fuel use differed in Mexico, where 88.45% of respondents used LNG or electricity, and was not associated with CRDs. Education was significantly associated with CRDs in both Ghana (p=0.0152) and Mexico (p=0.0123). In Ghana, age was significantly associated with CRDs, where, of those who had a CRDs, 49.91% were aged 65 or older, while among those who did not, only 33.28% were aged 65 or older.

### Model results

The ICC, calculated with a multilevel logistic regression model with no predictors except for the random intercepts for countries, was found to be below our predetermined cutoff (ICC: 0.205; 95%CI: 0.198-0.212), and subsequently, we proceeded with conventional logistic regression models to estimate the effects of cooking fuel on CRDs.

Table 2 presents the results for the unadjusted association between cooking fuel and the odds of having CRDs. In Ghana, those who used unclean cooking fuels had higher odds of having CRDs (OR: 2.93; 95%CI: 1.57-5.46).

**Table 2.**
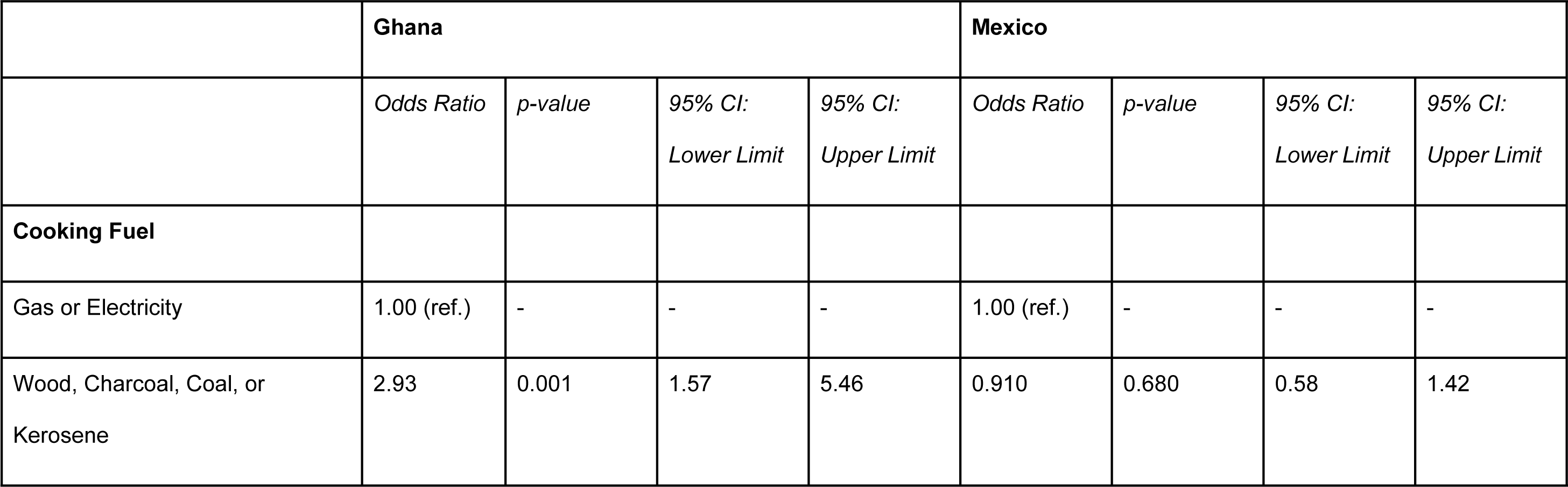
Unadjusted odds of reporting symptoms of chronic respiratory disease by primary household cooking fuel.

|  | Ghana |  |  |  | Mexico |  |  |  |
| --- | --- | --- | --- | --- | --- | --- | --- | --- |
|  | *Odds Ratio* | *p-value* | *95% CI:*<br>*Lower Limit* | *95% CI:*<br>*Upper Limit* | *Odds Ratio* | *p-value* | *95% CI:*<br>*Lower Limit* | *95% CI:*<br>*Upper Limit* |
| **Cooking Fuel** |  |  |  |  |  |  |  |  |
| Gas or Electricity | 1.00 (ref.) | - | - | - | 1.00 (ref.) | - | - | - |
| Wood, Charcoal, Coal, or<br>Kerosene | 2.93 | 0.001 | 1.57 | 5.46 | 0.910 | 0.680 | 0.58 | 1.42 |

This relationship continued to hold after adjusting for covariates (aOR: 2.74; 95%CI: 1.33-5.65) and are presented in Table 3. In both unadjusted and adjusted models, cooking fuel was not associated with differences in CRD odds among those living in Mexico. In both Ghana and Mexico, education was associated with differences in CRD risk. In Mexico, those who did not complete secondary education or who did not state their education attainment had higher odds of CRDs, the adjusted odds ratios were respectively 2.06 (95%CI: 1.34-3.18) and 1.96 (95%CI: 1.19-3.22). In Ghana, having less than secondary education was associated with odds (aOR: 0.54; 95%CI: 0.32-0.91).

**Table 3.** Adjusted odds of reporting symptoms of chronic respiratory disease by primary household cooking fuel. Stratified by country.

|  | **Ghana** |  |  |  | **Mexico** |  |  |  |
| --- | --- | --- | --- | --- | --- | --- | --- | --- |
|  | *Odds Ratio* | *p-value* | *95% CI:*<br>*Lower Limit* | *95% CI:*<br>*Upper Limit* | *Odds Ratio* | *p-value* | *95% CI:*<br>*Lower Limit* | *95% CI:*<br>*Upper Limit* |
| **Cooking Fuel** |  |  |  |  |  |  |  |  |
| Gas or Electricity | 1.00 (ref.) | - | - | - | 1.00 (ref.) | - | - | - |
| Wood, Charcoal, Coal, or<br>Kerosene | 2.74 | 0.007 | 1.33 | 5.65 | 1.02 | 0.934 | 0.60 | 1.75 |
| **Cooking Space** |  |  |  |  |  |  |  |  |
| Separate from Living/Sleeping<br>Space | 1.00 (ref.) | - | - | - | 1.00 (ref.) | - | - | - |
| Shared with Living/Sleeping Space | 4.06 | 0.018 | 1.28 | 12.85 | 5.44 | 0.029 | 1.19 | 24.76 |
| **Sex** |  |  |  |  |  |  |  |  |
| Male | 1.00 (ref.) | - | - | - | 1.00 (ref.) | - | - | - |
| Female | 0.97 | 0.864 | 0.66 | 1.42 | 0.90 | 0.517 | 0.67 | 1.23 |
| **Age** |  |  |  |  |  |  |  |  |
| Younger than 65 | 1.00 (ref.) | - | - | - | 1.00 (ref.) | - | - | - |
| 65 and Older | 1.87 | <0.0001 | 1.36 | 2.56 | 0.99 | 0.962 | 0.76 | 1.29 |
| **Education** |  |  |  |  |  |  |  |  |
| No Secondary Education | 0.54 | 0.020 | 0.32 | 0.91 | 2.06 | 0.001 | 1.34 | 3.18 |
| Secondary Education or Higher | 1.00 (ref.) | - | - | - | 1.00 (ref.) | - | - | - |
| Not Stated | 0.86 | 0.495 | 0.56 | 1.32 | 1.96 | 0.008 | 1.19 | 3.22 |
| **Rural-Urban Status** |  |  |  |  |  |  |  |  |
| Urban | 1.00 (ref.) | - | - | - | 1.00 (ref.) | - | - | - |
| Rural | 0.89 | 0.541 | 0.62 | 1.29 | 1.48 | 0.091 | 0.94 | 2.34 |
| **Smoking Status** |  |  |  |  |  |  |  |  |
| Never Smoked | 1.00 (ref.) | - | - | - | 1.00 (ref.) | - | - | - |
| Current Smoker | 1.54 | 0.192 | 0.80 | 2.97 | 0.84 | 0.410 | 0.54 | 1.28 |
| Former Smoker | 0.94 | 0.908 | 0.30 | 2.88 | 0.70 | 0.311 | 0.35 | 1.40 |
| Not Stated | 0.61 | 0.373 | 0.21 | 1.81 | 0.84 | 0.505 | 0.51 | 1.39 |
| **Hip-to-waist Ratio** | 1.66 | 0.469 | 0.42 | 6.65 | 0.91 | 0.194 | 0.78 | 1.05 |

Next, we conducted analyzed sex-stratified analyses for each country. In Ghana, females living in households that used solid fuels or kerosene had higher odds of CRDs (aOR: 2.60; 95%CI: 1.03-6.56). Females who cooked with solid fuels or kerosene and in a space shared with living/sleeping had a substantially higher risk for CRDs (aOR: 5.75; 95%CI: 1.04-31.82). Female current smokers had higher odds of CRDs (aOR: 7.79; 95%CI: 1.66-36.65). Males did not have significantly different risks by either smoking status, cooking fuel, or cooking space. Both females and males aged 65 and older had a higher risk, respectively having aORs 1.87 (95%CI: 1.36-2.56) and 1.86 (95%CI: 1.10-3.15). In Mexico, no significant differences in cooking fuel use were found for either females or males. As for cooking space, men living in households where solid fuels or kerosene was the primary cooking fuel and where the cooking space was shared with living/sleeping space had substantially higher odds of CRDs than those without (aOR: 9.39; 95%CI: 1.80-48.98). Women who did not complete secondary education and those who did not state their educational attainment both had higher odds of CRDs, respectively having adjusted odds ratios 2.51 (95%CI: 1.53-4.11) and 2.13 (95%CI: 1.17-3.90).

## Discussion

This study conducted a cross-sectional analysis using the Wave 2 data from the SAGE project, focusing on adults aged 50+ in Ghana and Mexico, to examine the association between cooking fuels and CRD symptoms. We found that in Ghana, individuals using solid fuels or kerosene had a significantly higher risk of CRDs compared to those using cleaner fuels like gas or electricity. This association was particularly pronounced among females and in households where cooking and living spaces were shared. In contrast, in Mexico, no significant association was found between cooking fuel type and CRD symptoms.

### Comparison with prior literature

While differing slightly in magnitude, the direction of the association between cooking fuel and CRD was similar to a prior Chinese study which found that solid fuels were associated with higher odds of CRDs (aOR: 1.30; 95%CI: 1.26-1.34). We note that this study differs from ours in that they only made comparisons between solid fuels and clean fuels (LNG, biogas, and electricity). As for gender differences, in Mexico, men living in households using solid cooking fuels and kerosene were associated with a higher risk of CRDs compared to women in similar settings. This finding contrasts to prior studies from other contexts that assign greater risks to women (58–60). A provisional explanation for this finding is that women might underreport their symptoms due to cultural norms or other societal factors, leading to an apparent lower risk in women when compared to men. Likewise, men may have additional exposure to respiratory irritants and pollutants through their occupational activities. If these exposures are more prevalent or intense than those experienced by women, they could contribute to the higher risk of CRDs observed in men.

### Strengths and limitations

There are some limitations associated with this study. First, due to its cross-sectional nature, the relationship between cooking fuel use and NCD is associational, and causality cannot be directly inferred from this study. However, as future waves of the SAGE study become available, longitudinal evidence may be obtained from the same population, which could help to establish temporal relationships and potentially infer causality. Second, the measure for CRD symptoms is subject to recall bias which may result in milder to moderate CRD symptoms being underreported, biasing our results towards the null.

This study boasts several notable strengths that reinforce the validity and importance of its findings. First, it leverages the comprehensive and standardized data collection framework of the SAGE study, ensuring consistency and reliability across a diverse sample. The large sample size, encompassing a broad demographic of older adults from Ghana and Mexico, enhances the generalizability of the results to similar contexts. Moreover, the inclusion of both self-reported symptoms and medical diagnoses in our outcome measure provides a comprehensive view of CRDs, balancing subjective and objective assessments. These methodological strengths collectively ensure that our study makes a significant contribution to the existing literature on environmental health and CRDs, particularly in the context of aging populations in low– and middle-income countries.

## Conclusion

In conclusion, this study provides critical insights into the relationship between household cooking fuel use and chronic respiratory diseases in older adults, particularly in the contexts of Ghana and Mexico. The findings are especially pertinent to public health policymakers and environmental health authorities, offering valuable evidence to guide interventions aimed at reducing the burden of CRDs. Our findings highlight the importance of increasing the accessibility of clean cooking methods (i.e., LPG and electricity) to reduce household air pollution and subsequent symptoms of CRDs. Healthcare providers can also benefit from these insights, as they provide an important perspective considering household environmental factors in the management and prevention of respiratory ailments. Ultimately, this study underscores the importance of addressing environmental determinants of health to improve outcomes for aging populations in LMICs.

## Data Availability

Data from SAGE Ghana Wave 2 & SAGE Mexico Wave 2 were used for this study. WHO SAGE was approved by the WHO Ethics Review Committee (reference number RPC149) with local approval from the University of Ghana Medical School Ethics and Protocol Review Committee (Ghana). The Ethics and Research Committees of the National Institute of Public health in Mexico approved the SAGE Mexico Study. All files were obtained from the World Health Organization Study on global AGEing and adult health (WHO SAGE). Details on data can be found at http://www.who.int/healthinfo/sage/cohorts/en/. The authors used files GhanaINDDataW2, GhanaHHDataW2, MexicoINDDataW2, and MexicoHHDataW2. The authors confirm that they had no special access privileges to the data. Interested researchers can apply to WHO to access data (http://apps.who.int/healthinfo/systems/surveydata/index.php/catalog/sage/about).

